# Intersectional factors associated with non-engagement in follow-up care among critical care survivors: A retrospective cohort study

**DOI:** 10.1101/2025.10.20.25338394

**Authors:** Saira Nazeer, Alexander J Fowler, Amar Ahmed, Tim Stephens, Yize Wan, Julie Sanders, John Prowle, Rupert Pearse, Zudin Puthucheary

**Author notes:** **<u>Declaration of interest:</u>** ZP has received honoraria for consultancy from GlaxoSmithKline, Lyric Pharmaceuticals, Faraday Pharmaceuticals and Fresenius-Kabi, educational support from Baxter and Nestle Health Science and speaker fees from Orion, Baxter, Sedana, Fresenius-Kabi and Nestle. RP undertakes paid consultancy work for Mode Sensors. SN and AJF have no relevant conflicts of interest. **Authors’ contributions** SN, TS, JS and ZP conceived and designed the study. SN, AA and AJF performed the analyses with input from JP, YW and RP. SN, AJF and ZP drafted the initial manuscript. All authors approved the final version of the manuscript.

## Abstract

**Introduction:** Recovery for those who have survived a critical illness can be a long process, with many survivors facing physical, psychological and cognitive challenges that can lead to long-term reliance on health and social care services. Engagement with follow up services is therefore essential for critical care survivors, however some face greater barriers to engagement leading to worse health outcomes. Understanding these disparities in service engagement is a crucial first step in equitable healthcare provision.

**Methods:** Retrospective cohort study including adults aged ≥16 years discharged from a critical care unit and who survived to hospital discharge between April 2023 and December 2024. Eligible patients with valid contact details were invited to complete an electronic questionnaire post-discharge from critical care. The primary outcome was engagement with a follow up service, defined as completion of the electronic questionnaire. Demographic and clinical data were obtained from routine local data collection. The primary exposures were index of multiple deprivation and ethnic group. Associations were determined with adjusted logistic regression. Intersectional analysis was conducted to understand relationships more comprehensively.

**Results:** A total of 2191 patients with complete data and who survived to hospital discharge were included in the analysis. 639 (29%) patients engaged with the follow up service and 1552 (71%) patients did not engage. There was no statistical significance in age, sex, or clinical characteristics between those who engaged and those who did not. Non-engagement was significantly associated with higher deprivation (aOR = 0.72, CI = 0.5 – 0.89), Black ethnicity (aOR = 0.60, CI = 0.44 – 0.80), and frailty (aOR = 0.72, CI = 0.50 – 0.89). Intersectional analysis revealed subgroups with high rates of non-engagement, individuals of Asian ethnicity from less deprived areas.

**Conclusions:** The method of analysing disparities and factors for non-engagement can significantly affect which populations are revealed to be at risk. Applying an intersectional lens is essential for identifying groups who may be overlooked by single factor approaches. Understanding the size of these at-risk groups, rather than just their relative odds ratios, is vital for planning interventions to maximise impact. Failure to account for intersectionality risks designing interventions that are ineffective or inequitable, ultimately perpetuating existing disparities in recovery outcomes for critical care survivors.

## Introduction

Over 200,000 people are admitted to intensive care units (ICU) in the United Kingdom annually (1). 70% survive to discharge, but over half experience hospital readmission within a year and many develop new chronic conditions (2–9). These physical, psychological and cognitive challenges, collectively termed Post Intensive Care Syndrome (PICS), can lead to long-term reliance on health and social care services (4,10–12). National guidelines recommend follow-up care to address these complex needs (13,14). However follow-up service engagement is variable (15,16). Failure to address patient engagement contributes to persistent health inequalities, poorer health outcomes, and increased demand on health and social care services, resulting in significant personal, societal and economic costs (17).

Healthcare policies and quality improvement initiatives increasingly recognise patient engagement as a critical factor influencing healthcare utilisation and outcomes (18,19). Policy makers have highlighted the importance of supporting patient engagement to improve service uptake and adherence and to reduce the impact of social determinants of health (18–21). Patient engagement relies on the coordinated partnerships between patients and healthcare providers (22). However service engagement is not shaped by a single factor but by the dynamic interplay of multiple social identities and structural constructs – a concept known as intersectionality (23). Ethnicity and socio-economic status have emerged as important factors influencing patient engagement, with ethnic minority (term used for consistency with existing literature, though “global majority” is acknowledged as more accurate and inclusive) and socio-economically deprived groups often facing greater barriers to engagement and worse health outcomes (24–27). Conventional determinants of causal and associative factors may be too simplistic as intersectionality is a framework determined by the overlap of identity factors (such as race, gender, socio-economic status and disability) with systems of power (such as racism, classism and ableism), interacting to shape experiences (23,28,29). Social determinants of health and service accessibility are deeply intertwined with these intersectional identities and are key drivers of health disparities and engagement barriers (24,30,31).

Identifying factors of non-engagement therefore requires acknowledgement of these intersecting factors and careful consideration when selecting analysis methodologies, so as to ensure we are truly investigating the complex intersecting nature of non-engagement, rather than simply revealing single factor relationships. We examined our population of critical care survivors for features of non-engagement. We hypothesised that there would be an association between an individual’s socioeconomic status and ethnicity, and their ability to engage with a critical care follow up service. We aimed to identify subgroups of patients who did not engage.

## Methods

### Study design

We performed a retrospective cohort study of patients admitted to the Adult Critical Care Unit at the Royal London Hospital, UK, a large unit serving a mixed patient population including trauma and neurocritical care patients, between April 2023 and December 2024. Patients aged 16 years and older who survived to hospital discharge and for whom complete data sets were available were included in the analysis. We excluded those with missing Acute Physiology and Chronic Health Evaluation (APACHE), Sequential Organ Failure Assessment (SOFA), Indices of Multiple Deprivation (IMD) and frailty scores. We excluded patients who did not have valid contact details (phone number and/or email address) recorded in hospital records. Research ethics approval was obtained from the Health and Care Research Ethics Committee (reference number 23/YH/0146, 22.08.2023). We report our findings in line with the STROBE checklist for cohort studies (32).

### Exposures

Data were collected as part of our Intensive Care National Audit and Research Centre (ICNARC) routine data collection. Demographic data included age on admission in whole years (owing to the small number of patients at either end of the age range, age was grouped into ‘≤17 years’ and ‘≥ 90 years’), sex (at birth, as recorded in medical records), and ethnicity (grouped by Office of National Statistics census categories into Black, Asian, Mixed/Other, White and Not Stated). We determined socio-economic status from the recorded postcode, which we then linked to Office for National Statistics lower super output area to obtain the index of multiple deprivation (IMD). We grouped IMD into quintiles based on the IMD rank. Clinical data included: level of frailty (Clinical Frailty Score in conjunction with clinician based assessment informed by the subjective history documented in the medical notes – owing to the small number of patients classified as having a ‘stable long term condition’, this category was combined with the ‘frail’ category for analytical purposes), admitting speciality (grouped into medical, surgical, neurosurgical and trauma), critical care length of stay (LOS), maximum organ support, Acute Physiology and Chronic Health Evaluation (APACHE) and the Sequential Organ Failure Assessment (SOFA) score. The primary exposures of interest were ethnicity and IMD.

### Data collection

Patients were sent an electronic questionnaire 72 hours post-discharge from critical care, with two reminder messages sent in the following 48 hours and then a new set sent 4 weeks post discharge from critical care. The intention of the questionnaire was to assist in identifying needs post critical illness and therefore consisted of a combination of four quality of life patient related outcome measures, selected to cover the vast range of symptoms an individual may experience post critical illness. It was available in two additional languages (Polish and Bengali) selected to reflect the local population.

### Outcome measures

The primary outcome was engagement. Engagement was defined as completion of part or all of the questionnaire at any time. For the purpose of this study, ‘non-engagement’ refers to when a patient did not complete any of the questionnaire.

### Statistical analysis

Descriptive statistics were used to summarise characteristics of the cohort. Associations between characteristics and engagement status were assessed using Chi-squared tests for categorical variables and Mann-Whitney U tests for continuous variables. We used logistic regression for univariate analyses with engagement as the dependent variable. All variables apart from the SOFA score were included in a multivariable logistic regression. We present odds ratios with associated 95% confidence intervals. To explore the interaction between intersectional features, we examined the distribution of engagement rates across groups defined by IMD quintiles and ethnicity, and compared counts and engagement rates within these intersectional groups. Furthermore, we included an interaction term between ethnicity and IMD quintiles in a multivariable logistic regression model. All analyses were performed using R software (R software foundation, Vienna, version 4.4.3) and associated packages ggplot2 (ref).

## Results

### Study population

From a total of 3058 critical care survivors, 2191 had complete data sets and survived to hospital discharge (figure 1). These data sets made up the cohort for subsequent analysis. The overall cohort of critical care survivors, had an average age of 53 (39-67), were predominantly male (1382/2191, 63%) and of White ethnicity (1101/2191, 50%), living in areas of high socioeconomic deprivation (1216/2191, 56% from the lowest 3 IMD deciles) and were admitted under medical (800/2191, 36%), neurosurgical (301/2191, 14%), surgical (652/2191, 30%) and trauma specialities (438/2191, 20%). We describe the characteristics and outcomes of patients with missing data (supplementary material table 1).

**Table 1:** Patient characteristics for overall cohort (n=2191), stratified for engagement. Numbers are presented as n (% of total) unless otherwise stated. To calculate p values, Mann-Whitney U was used for continuous data and chi-squared tests for categorical data. Data regarding residence prior to admission has been omitted for data integrity concerns. IMD quintiles based on IMD rank within the cohort.

|  | Engagement Status |  |  |
| --- | --- | --- | --- |
|  | Non-Engaged<br>(n=1552) | Engaged<br>(n=639) | p-value |
| **AGE** | 56 (39 to 68) | 54 (39 to 66) | 0.14 |
| **ETHNICITY** |  |  |  |
| Asian | 360 (23.2%) | 136 (21.3%) | <0.05 |
| Black | 266 (17.1%) | 74 (11.6%) | <0.05 |
| Mixed/Other | 69 (4.4%) | 33 (5.2%) | <0.05 |
| Not stated | 103 (6.6%) | 49 (7.7%) | <0.05 |
| White | 754 (48.6%) | 347 (54.3%) | <0.05 |
| **SEX** |  |  |  |
| Female | 565 (36.4%) | 244 (38.2%) | 0.46 |
| Male | 987 (63.6%) | 395 (61.8%) | 0.46 |
| **IMD QUINTILES** |  |  |  |
| 1 - most deprived | 326 (21%) | 112 (17.5%) | <0.05 |
| 2 | 327 (21.1%) | 111 (17.4%) | <0.05 |
| 3 | 316 (20.4%) | 122 (19.1%) | <0.05 |
| 4 | 293 (18.9%) | 145 (22.7%) | <0.05 |
| 5 - least deprived | 289 (18.6%) | 149 (23.3%) | <0.05 |
| **FRAILITY** |  |  |  |
| Frail | 272 (17.5%) | 76 (11.9%) | <0.05 |
| Not frail | 1280 (82.5%) | 563 (88.1%) | <0.05 |
| **MAX ORGAN SUPPORT** |  |  |  |
| 0 | 29 (1.9%) | 12 (1.9%) | 0.47 |
| 1 | 816 (52.6%) | 346 (54.1%) | 0.47 |
| 2 | 489 (31.5%) | 200 (31.3%) | 0.47 |
| 3 | 201 (13%) | 79 (12.4%) | 0.47 |
| 4 | 17 (1.1%) | 2 (0.3%) | 0.47 |
| **ADMITTING SPECIALITY** |  |  |  |
| Medical | 582 (37.5%) | 218 (34.1%) | <0.05 |
| Neurosurgery | 219 (14.1%) | 82 (12.8%) | <0.05 |
| Surgical | 427 (27.5%) | 225 (35.2%) | <0.05 |
| Trauma | 324 (20.9%) | 114 (17.8%) | <0.05 |
| **SOFA SCORE** | 4 (2 to 5) | 3 (2 to 5) | <0.05 |
| **APACHE SCORE** | 13 (10 to 17) | 12 (9 to 16) | <0.05 |
| **CRITICAL CARE LOS** | 6 (4 to 11) | 6 (4 to 10) | 0.12 |

**APACHE** Acute Physiology and Chronic Health Evaluation, **IMD** Indices of Multiple Deprivation, **LOS** Length of Stay, **SOFA** Sequential Organ Failure Assessment

**Figure 1:**
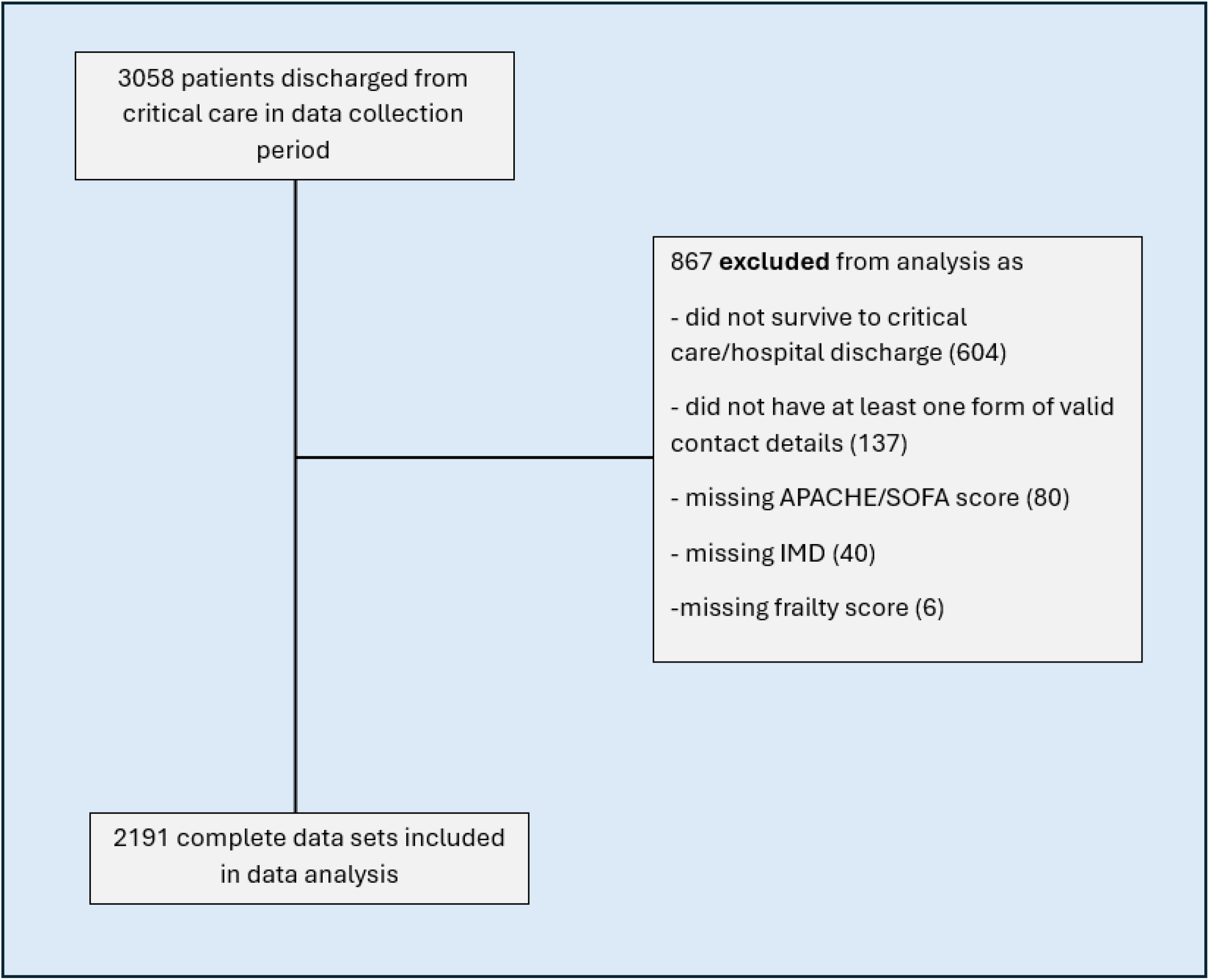
Flow diagram describing those we excluded from subsequent data analysis. Summary of incomplete data sets can be found in supplementary material. *APACHE* Acute Physiology and Chronic Health Evaluation, *IMD* Indices of Multiple Deprivation, *SOFA* Sequential Organ Failure Assessment

### Patient outcomes

A large proportion of the patients (1558, 71%) did not engage with the follow up service (Table 1). Median age and sex distribution did not differ significantly between groups, however ethnicity and socioeconomic status (measured by IMD) did significantly differ. Univariate logistic regression was first performed to assess associations between individual demographic and clinical characteristics and engagement status. Patients of Black ethnicity had significantly lower odds of engagement compared to those of White ethnicity (OR = 0.6 (0.45-0.8), p = <0.05). This association remained significant after adjusting for age, sex, clinical frailty score, admission speciality, and illness severity in the multivariable logistic regression model (aOR = 0.6 (0.44-0.8), p = <0.05). Similarly, patients from more socioeconomically deprived areas had lower odds of engagement. Compared to those in the least deprived IMD quintile, individuals in more deprived quintiles had significantly lower odds of engagement (OR = 0.67 (CI = 0.5-0.89), p = <0.05). This relationship also remained significant in the multivariable model (aOR = 0.72 (0.53-0.98), p = <0.05) (Table 2, Figure 2).

**Table 2:** Logistic regression analysis. Numbers are presented as OR (CI) or the p value. For categorical data, the reference category is stated in the table.

| **VARIABLE** | **UNIVARIATE ANALYSIS** |  | **MULTIVARIABLE ANALYSIS** |  |
| --- | --- | --- | --- | --- |
|  | **Unadjusted OR** | **p value** | **Adjusted OR** | **p value** |
| **AGE** | 0.96 (0.91-1.01) | 0.13 | 0.96 (0.9-1.02) | 0.19 |
| **SEX** |  |  |  |  |
| Male | 0.93 (0.77-1.12) | 0.43 | 0.83 (0.68-1.01) | 0.07 |
| Female | Reference category |  |  |  |
| **ETHNICITY** |  |  |  |  |
| Asian | 0.82 (0.65-1.04) | 0.1 | 0.81 (0.63-1.03) | 0.09 |
| Black | 0.6 (0.45-0.8) | <0.05 | 0.6 (0.44-0.8) | <0.05 |
| Mixed / Other | 1.04 (0.67-1.59) | 0.86 | 1.01 (0.65-1.57) | 0.97 |
| Not stated | 1.03 (0.71-1.48) | 0.86 | 1.01 (0.69-1.46) | 0.98 |
| White | Reference category |  |  |  |
| **IMD QUINTILE** |  |  |  |  |
| 1 – most deprived | 0.67 (0.5-0.89) | <0.05 | 0.72 (0.53-0.98) | <0.05 |
| 2 | 0.66 (0.49-0.88) | <0.05 | 0.68 (0.5-0.92) | <0.05 |
| 3 | 0.75 (0.56-1) | <0.05 | 0.8 (0.59-1.07) | 0.13 |
| 4 | 0.96 (0.72-1.27) | 0.77 | 1 (0.75-1.34) | 0.98 |
| 5 – least deprived | Reference category |  |  |  |
| **MAX ORGAN SUPPORT** | 0.94 (0.83-1.06) | 0.3 | 1.04 (0.9-1.2) | 0.63 |
| **APACHE II SCORE** | 0.98 (0.96-0.99) | <0.05 | 0.98 (0.96-1) | 0.11 |
| **FRAILITY** |  |  |  |  |
| Frail | 0.64 (0.48-0.83) | <0.05 | 0.66 (0.5-0.89) | <0.05 |
| Not frail | Reference category |  |  |  |
| **CRITICAL CARE LOS (days)** | 0.99 (0.98-1) | <0.05 | 0.99 (0.98-1) | 0.06 |
| **ADMITTING SPECIALITY** |  |  |  |  |
| Neurosurgery | 1 (0.74-1.34) | 1 | 0.89 (0.64-1.22) | 0.46 |
| Surgical | 1.41 (1.12-1.76) | <0.05 | 1.33 (1.05-1.68) | <0.05 |
| Trauma | 0.94 (0.72-1.22) | 0.64 | 0.78 (0.59-1.04) | 0.09 |
| Medical | Reference category |  |  |  |

**Figure 2:**
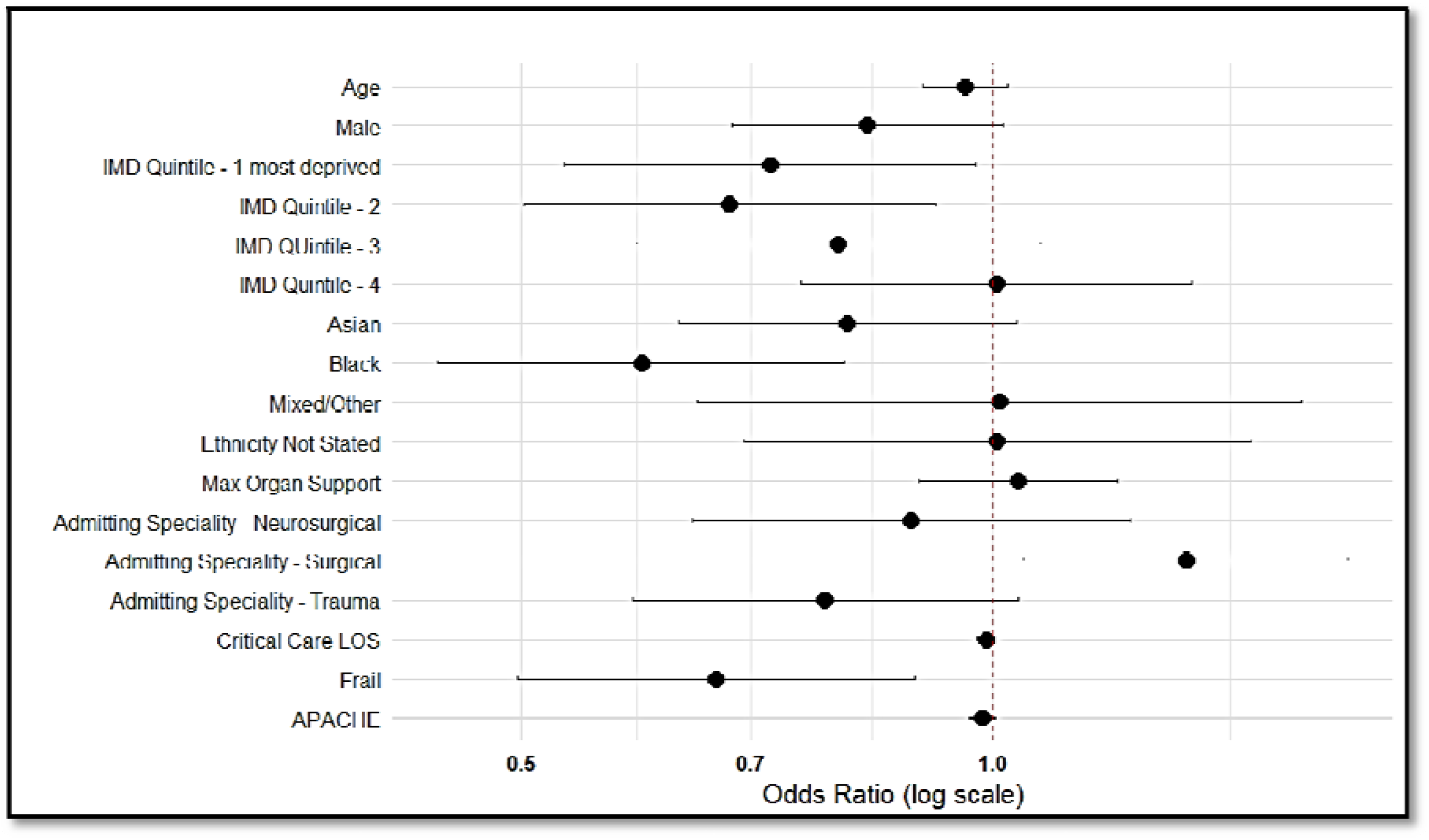
Forest plot depicting the odds ratios for factors of non-engagement. Adjusted odds ratios for all variables used in multivariate logistic regression analysis. Reference categories same as shown in Table 3. An odds ratio of <1 shows factors associated with non-engagement (i.e. that make you more at risk of non-engagement), whereas an odds ratio of >1 shows factors associated with engagement. ***APACHE* Acute Physiology and Chronic Health Evaluation, *IMD* Indices of Multiple Deprivation, *LOS* Length of Stay**

Further analysis of our data was conducted to understand the intersecting impact of socioeconomic status and ethnicity on an individual’s ability to engage. The introduction of an interaction term to the logistic model made no significant difference to the results. A heat map (Table 3) allowed us to distinguish patterns of non-engagement. We could clearly see the groups revealed by the traditional single factor analyses: IMD quintiles 1 and 2 and Black ethnicity groups, including subgroups of interest within these - a small group of individuals from Black ethnic backgrounds living in the least deprived areas and a large group of White individuals living in areas of high deprivation. However, this analysis also revealed subgroups of interest outside of the categories defined by the single factor analyses; with the most obvious being a large group of Asian ethnicity individuals from mid to low socioeconomic deprivation (IMD quintiles 3 and 4) displaying poor engagement. This heat map additionally highlighted the size of populations allowing this to be a factor to consider when analysing the data. Factors revealed through traditional analysis (IMD quintiles 1 and 2 and Black ethnicity) identify 1,039 critical care survivors at risk of non-engagement, whereas this more nuanced intersectional approach reveals a further 233.

**Table 3:**
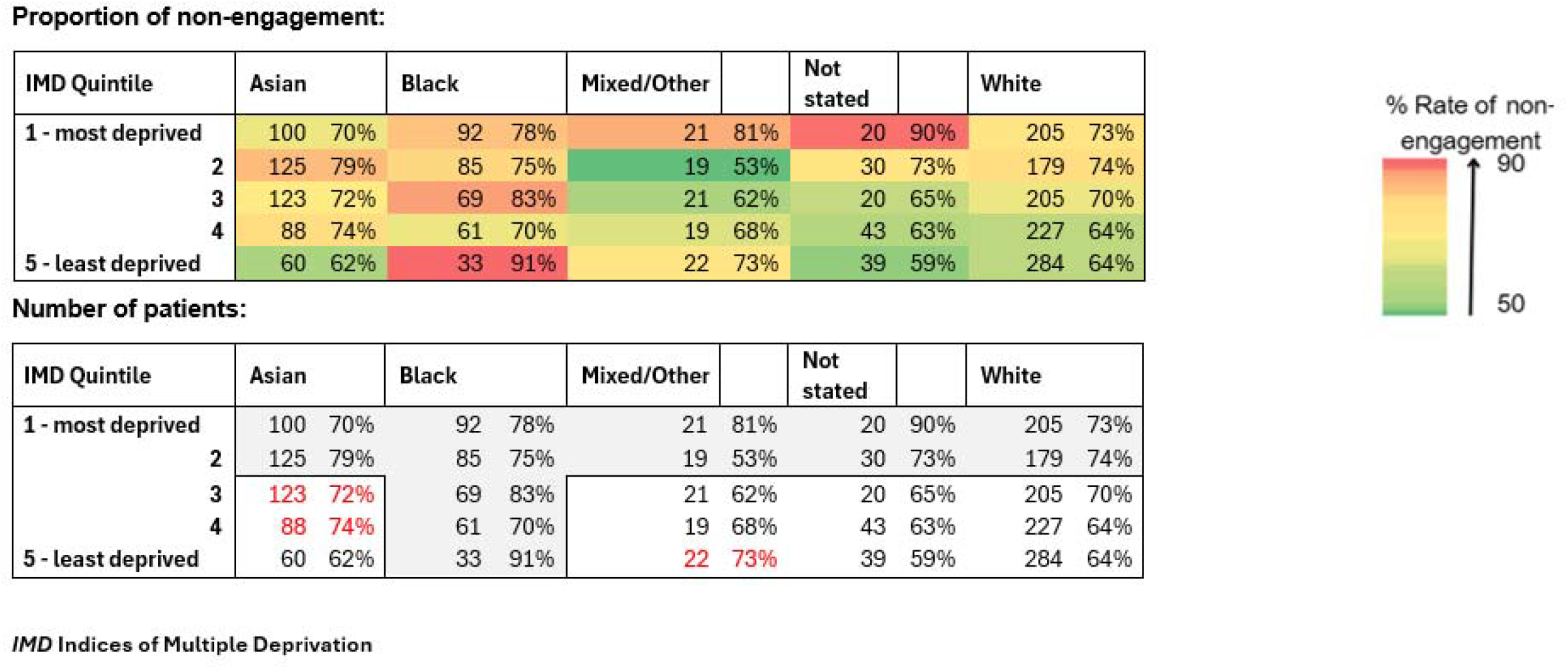
A visualisation of the intersectionality of engagement. The first table heat maps the percentage of non-engagement with the follow-up service in our cohort by ethnicity and socioeconomic status. n= number of people in each group, and the colour represents the non-engagement rate in the critical care follow up service (see scale). The second table depicts the same interaction, whilst also highlighting the subgroups (in red) with high levels of non-engagement (>71% - the average non-engagement rate across the total cohort) overlooked by single factor analysis (highlighted box). These tables help visualise the groups at risk of non-engagement, alongside the scale of that risk.

## Discussion

We demonstrate that using different methodologies reveals different factors and populations associated with non-engagement with a critical care follow up service. Patients from more socioeconomically deprived areas and those from Black ethnic backgrounds had consistently higher odds of non-engagement, even when adjusting for confounders. However when an intersectional analysis was applied, additional patterns of non-engagement that were not apparent through single factor approaches were seen. Our analyses highlight the risk that traditional logistic regression analysis may overlook the scale of the problem by obscuring large groups of individuals who would additionally benefit from focussed/tailored interventions. These insights underscore the value of intersectional methods in uncovering both visible and obscured dimensions of inequity in access and engagement with healthcare.

Our findings largely align with existing literature, which describes lower engagement in ethnic minority groups due to barriers such as language, lower health literacy, and cultural mismatch (30,33,34). Lower activation and engagement correlate with worse health outcomes, including higher readmission rates, increased morbidity and reduced quality of life (35). Additionally there is the known impact of socio-economic disadvantage on healthcare outcomes and access for critical care survivors (27,36,37). However, findings of subgroups of individuals of Black ethnicity in the least deprived areas showing the highest levels of non-engagement, alongside a large group of Asian individuals from IMD quintiles 3 and 4 are less well described, and require further investigation.

## Strengths and limitations

A key strength of this study is the use of intersectional visualisation techniques, which allowed for a more detailed exploration of non-engagement patterns across social dimensions that are often examined in isolation. This approach helped reveal both hidden small groups at risk and large populations of unmet needs, offering novel insights to inform policy and practice. The diversity of the study population facilitated this comprehensive analysis, revealing inequities that may not have been apparent in less heterogenous cohorts. Although those without valid contact details were excluded from data analysis, reliance on electronic communication may have disadvantaged individuals experiencing digital poverty and/or poor digital literacy. However digital poverty was unlikely to be a major factor given the small number excluded (137, 4% of the total population), but digital literacy remains a potentially important and underexplored barrier in this population. Comparable services within the same NHS Trust have achieved engagement rates of up to 60-80%, suggesting that digital exclusion alone does not fully explain the lower response rate observed in our population. Data integrity and inaccuracies may have introduced a source of bias and influenced the findings. Additionally, as this questionnaire was distributed via an automated service to a population which is heterogenous in it’s recovery trajectory and timeline, it is likely that for some it did not coincide with a point at which they were clinically able to complete it or considered it relevant. To mitigate this, reminder messages were sent including a second distribution four weeks later. Finally, as this study was conducted in a single large inner-city trauma centre, the results may not be fully generalisable to other settings with different population profiles or service structures.

### Implications for future practice

Our findings emphasise the need for tailored engagement strategies that account for intersecting identities and structural barriers, rather than focussing on single factors in isolation. Intervention developers must recognise that disadvantage rarely exists in a vacuum, but rather through the compounding intersecting effects of multiple factors. Embedding intersectionality into intervention design is therefore essential to ensure that engagement strategies meet the complex needs of critical care survivors and reduce inequities in access.

### Implications for future research

Routinely collected hospital data fail to capture many key factors of structural predictors of non-engagement. Evidence suggests that greater social support is associated with higher patient activation and improved engagement (38). Components of social support such as informal care arrangements and insecure housing will affect an individual’s ability to engage, however they are poorly recorded or absent from hospital datasets. Linkage to social care records will provide some of the answers, however a comprehensive understanding of engagement in this population will require prospective data collection to ensure all relevant determinants are considered. Future research will need to employ codesign methodologies to ensure relevance and responsiveness to the needs of critical care survivors and their communities. A caveat is that co-design processes often attract individuals who are already engaged, which risks overlooking the perspectives of those less likely to participate. Using pre-existing trusted community pathways and networks to reach and involve groups who might otherwise remain disengaged may mitigate this. Local faith groups, community centres, peer support networks, or voluntary organisations with established relationships can help build trust and facilitate access to underrepresented groups. It is also important to note, that studies evaluating tailored engagement interventions in other areas of healthcare rarely achieve universal engagement (39–42). This indicates that a more nuanced approach may be required, one that considers multiple intersecting factors when designing engagement strategies.

## Conclusions

The method of analysing disparities and risk factors for non-engagement can significantly affect which populations are revealed to be at risk. Applying an intersectional lens is essential for identifying groups who may be overlooked by single factor approaches. Additionally, understanding the size of these groups is vital in the planning of interventions to maximise impact. Failure to account for intersectionality risks designing interventions that are ineffective or inequitable, ultimately perpetuating existing disparities in recovery outcomes for critical care survivors.

## Data Availability

Data can be made available upon reasonable request to corresponding author.

## Acknowledgments

We would like to thank the ICNARC team at The Royal London Hospital for collecting such rich and valuable data.

